# Higher clinical acuity and 7-day hospital mortality in non-COVID-19 acute medical admissions: prospective observational study

**DOI:** 10.1101/2020.06.26.20098434

**Authors:** Marcus J Lyall, Nazir I Lone

## Abstract

**Objectives:** To understand the effect of COVID-19 lockdown measures on severity of illness and mortality in non-COVID-19 acute medical admissions.

**Design:** A prospective observational study

**Setting:** 3 large acute medical receiving units in NHS Lothian, Scotland. Participants: Non-covid-19 acute admissions (n = 1756) were examined over the first 31 days after the implementation of the COVID-19 lockdown policy in the United Kingdom on 23^rd^ March 2019. Patients admitted over a matched interval in the previous 5 years were used as a comparator cohort (n = 14961).

**Main outcome measures:** Patient demography, biochemical markers of clinical acuity and 7-day hospital inpatient mortality.

**Results:** Non-covid-19 acute medical admissions reduced by a mean 43.8% (95% CI 27.3, 59.4) across all 3 sites in comparison to the mean of the preceding 5 years P < 0.001. The reduction in admissions predominated in the over 75 age category and a greater proportion arrived by emergency ambulance transport. Non-covid-19 admissions during lockdown had a greater incidence of severe renal injury, hyperlactataemia and over twice the risk of hospital death within 7 days 5.01% vs 2.49% which persisted after adjustment for confounders (OR 2.17, 95% CI 1.70,2.73, P < 0.0001)

**Conclusions:** These data support current fears that patients are delaying seeking medical attention for acute illness which is associated with worsening clinical parameters and a higher risk of death following admission.

## Introduction

COVID-19, the disease manifestation of the SARS-CoV-2 virus global pandemic in 2020 has enforced unprecedented change on how the people of the United Kingdom live their lives. From 23^rd^ March 2020 measures were taken to slow the spread of SARS-CoV-2 with the closure of entertainment, hospitality and indoor leisure premises, the advice to stay home and limit all but essential travel and to work from home where at all possible [1].

Self-attendance rates to emergency medicine services have sharply declined, with a marked reduction in patients presenting with a possible myocardial infarction prompting concern that patients with significant acute illness may not be attending hospital for acute medical care [2]. The reasons for this are unclear but may be due to concerns about being infected with SARS-CoV-2 or burdening the health service during the pandemic. Concerns that this has resulted in public harm are supported by the most recent data from the from the Office of National Statistics UK, recording deaths of 22351 for week 16 in 2020 in England and Wales, 11854 more than the five-year average for this week. While COVID-19 is listed on the death certificate in 8758 deaths, this does not represent all of the excess [3]. Similar concerns have been raised following the weekly data report from the National Records of Scotland demonstrating a 79% increase in all-cause mortality for the same week, with 23% of the excess mortality not attributed to COVID-19 [4].

Community testing for presumed SARS-CoV-2 infection was not adopted in this period. As such it is unclear whether a proportion of this excess mortality results from delayed presentation to health care services with potentially life-threatening conditions unrelated to SARS-CoV-2 infection. To investigate this further, we prospectively examined the demography, route of admission, blood markers of medical acuity and adjusted hospital 7- day mortality of non-COVID-19 acute medical admissions to three large acute medical units in a health board in Scotland during the lock down period. Admissions to the same units over the same time frame in the proceeding 5 years were used as comparison.

## Methods

Data were obtained from a Trakcare inpatient management system (Intersystems, Illinois) and analysed using R version 3.6.1. Patients resident in the NHS Lothian Health Board area who were admitted to the three acute medical units during the 31 day period were examined following the first week of lockdown (23/03/2020). This was compared with the same 31 day period beginning the same year week from the preceding 5 years (13^th^ week of the year 2015-2019). Scottish Index of Multiple Deprivation (SIMD) quintile was obtained from the SIMD database 2020 version published by the Scottish government, population estimates were obtained from the National Records of Scotland database. Clinical laboratory test results provided measurements for blood lactate, serum creatinine and SARS-CoV-2 test results. Baseline creatinine was obtained from the year prior to the patient’s admission and Acute Kidney Injury network (AKIN) score [5] determined on the most recent blood test at least 7 days from hospital admission. Where baseline creatinine was not available, the median value of the population without known renal impairment was used (69umol/l). All admitted patients were screened for signs and symptoms of SARS-CoV-2 infection using local guidelines (supplementary material 1) and tested using rtPCR nose and throat combined swab where criteria are met. If this was negative and clinical suspicion persisted a second swab was performed. Any patients testing positive at any stage during admission were removed from the study. Admission rates were calculated for the lockdown period relative to the mean of the previous 5 years and P value determined using Poisson regression. 7-day hospital mortality was compared for the lockdown period relative to previous years using binary logistic regression and adjusted for age, sex and deprivation. Graphical outputs were performed using ggplot2 package. The study was reviewed by the Quality Improvement Team and registered in NHS Lothian as a Quality Improvement project. Following the HRA decision tool and after seeking advice from NHS Lothian Research and Development department, the study was deemed to be service evaluation and therefore formal ethical approval was not required. All data were anonymised before analysis and complied with local data protection requirements.

## Results

1776 non-COVID-19 medical admissions were identified during the 2020 lockdown period and compared to 14377 acute medical admissions from a matched period in the previous 5 years. Non-COVID-19 admissions to acute medical units fell by a mean of 43.8% (95% CI 27.3, 59.4) at the three hospital sites in comparison to the mean of the preceding 5 years (P < 0.001, figure 1). There was a reduction in the proportion of admitted patients aged over 75 whilst the proportion of patients in categories 25 years to 75 years were either unchanged or increased compared to previous years (figure 2). The source of attendance for patients changed with comparatively more attending through 999 emergency ambulance and fewer through NHS 24 recommendation or self-referral to the emergency department (figure 3). When examining acuity of illness, patients admitted in the lock down period had a higher prevalence of severe acute kidney injury and elevated serum lactate compared to the preceding 5 years (figures 4 + 5). We went on to examine the risk of 7-day in hospital mortality for the lockdown period in comparison to previous years. In keeping with the higher acuity of illness, patients admitted to acute medical units during the COVID-19 lockdown period had over twice the risk of death when compared to the previous 5 years 5.01% vs 2.49% which persisted after adjustment for confounders (OR 2.17, 95% CI 1.70,2.73, P < 0.0001)(figure 6).

**Figure 1.**
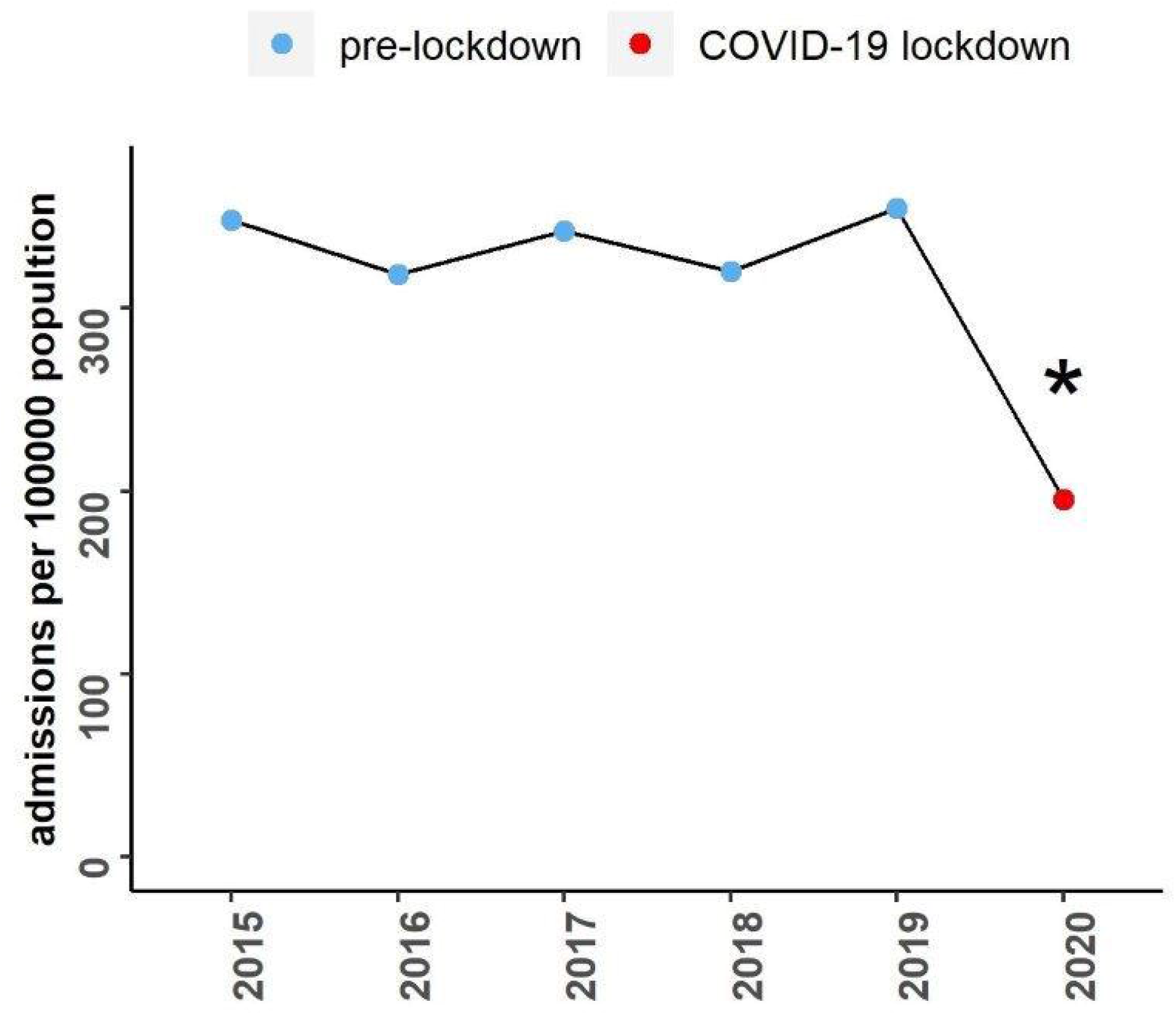
total admissions.

**Figure 2.**
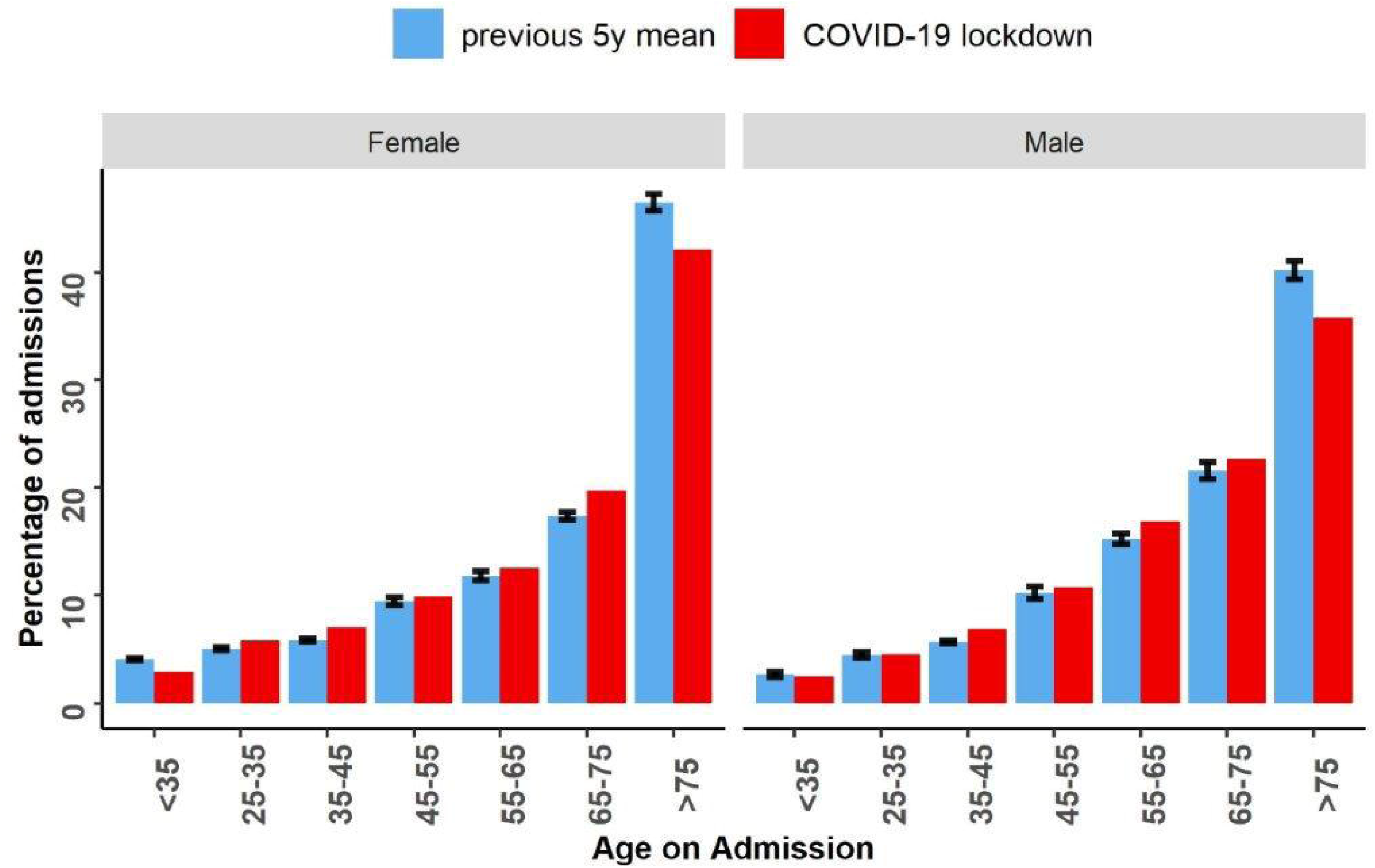
age group.

**Figure 3.**
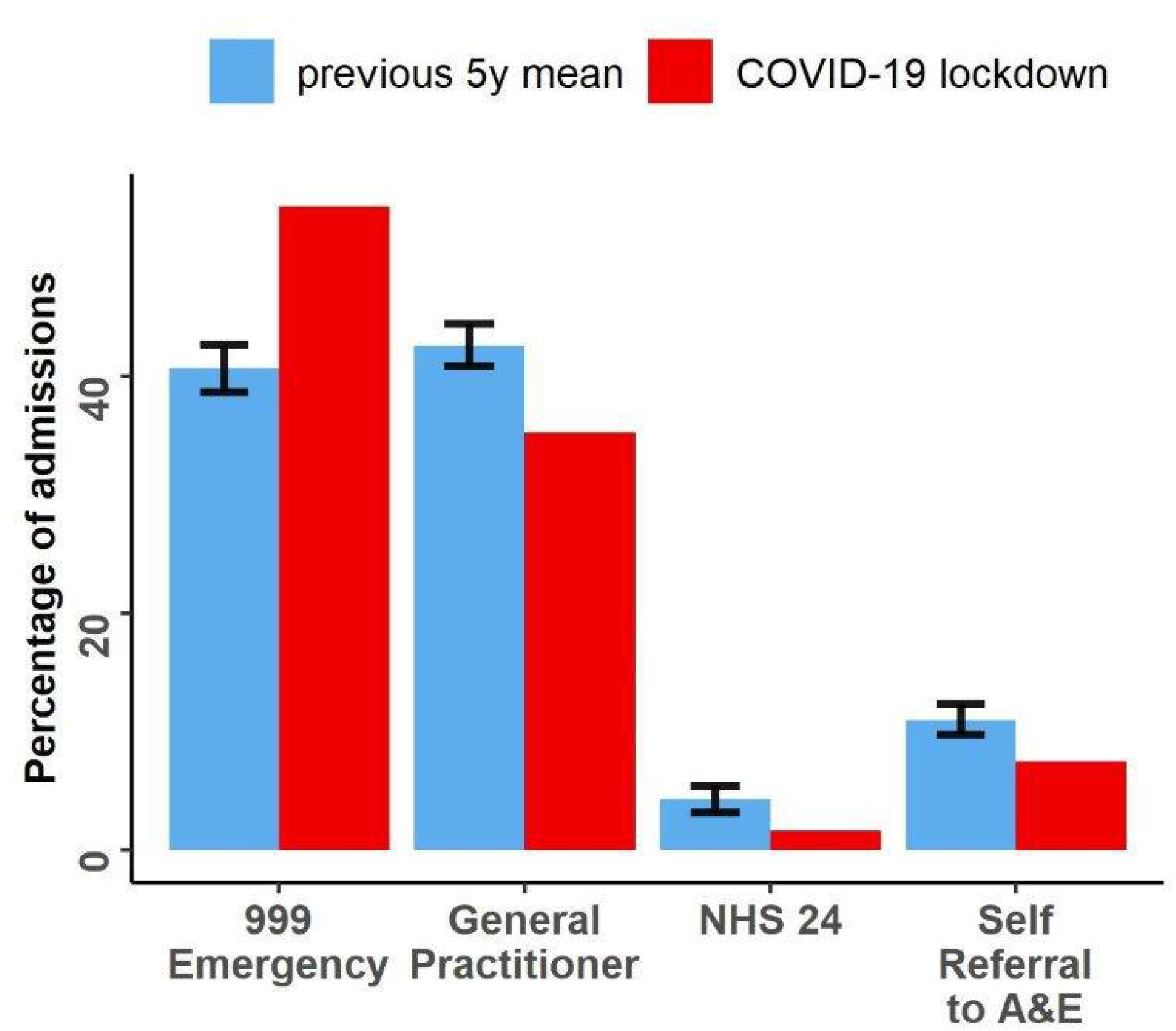
admission source to acute medical units during COVID-19 lockdown period versus last 5 years. (pre-lockdown n = 14961, COVID-19 lockdown n = 1776). * = P < 0.001 Poisson regression versus all previous years. error bars = +/- standard error.

**Figure 4.**
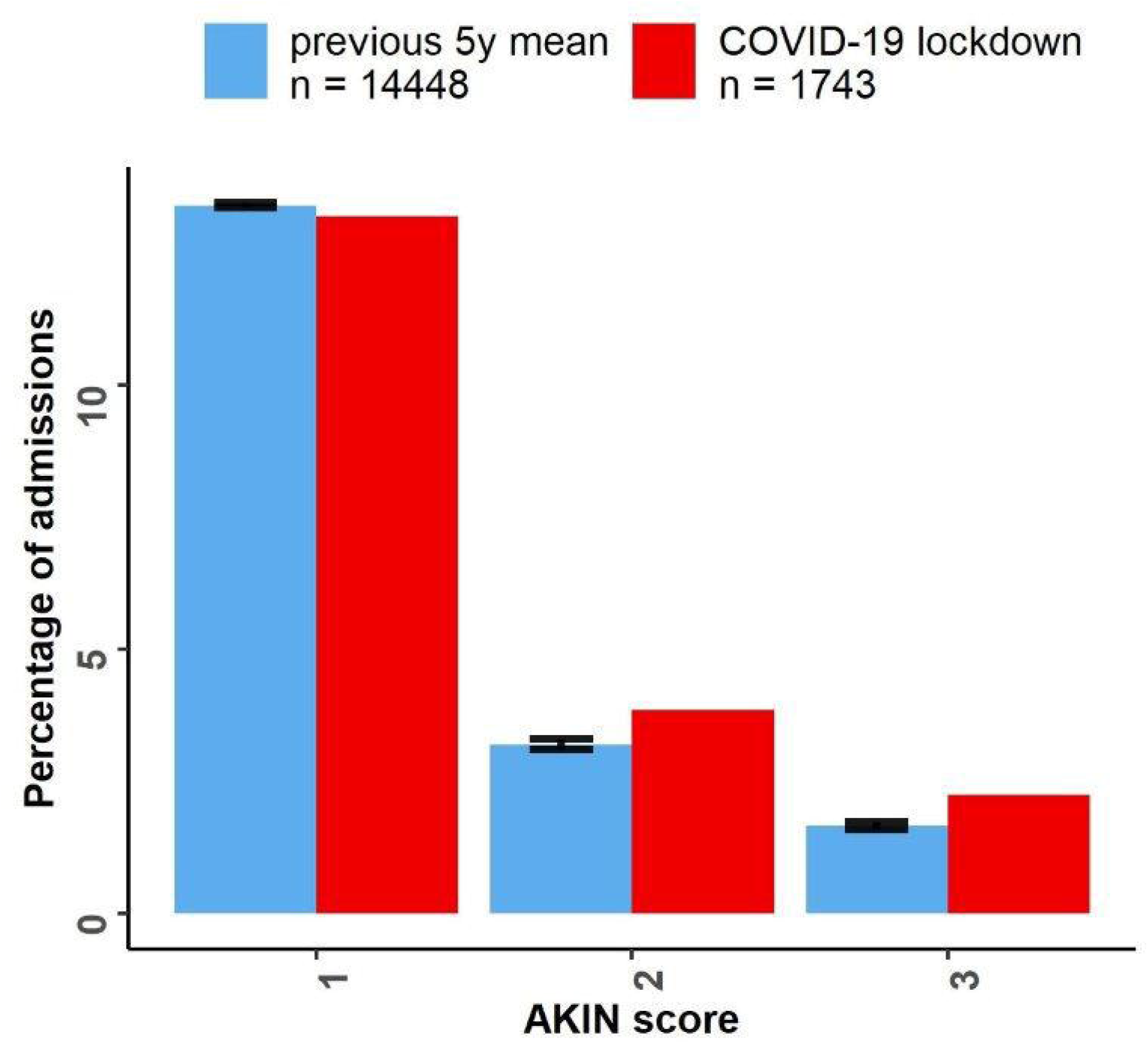
Proportion of admissions with AKIN stages of acute kidney injury in non-COVID-19 admissions in increasing severity from mild (1) to severe (3). error bars = +/- standard error.

**Figure 5.**
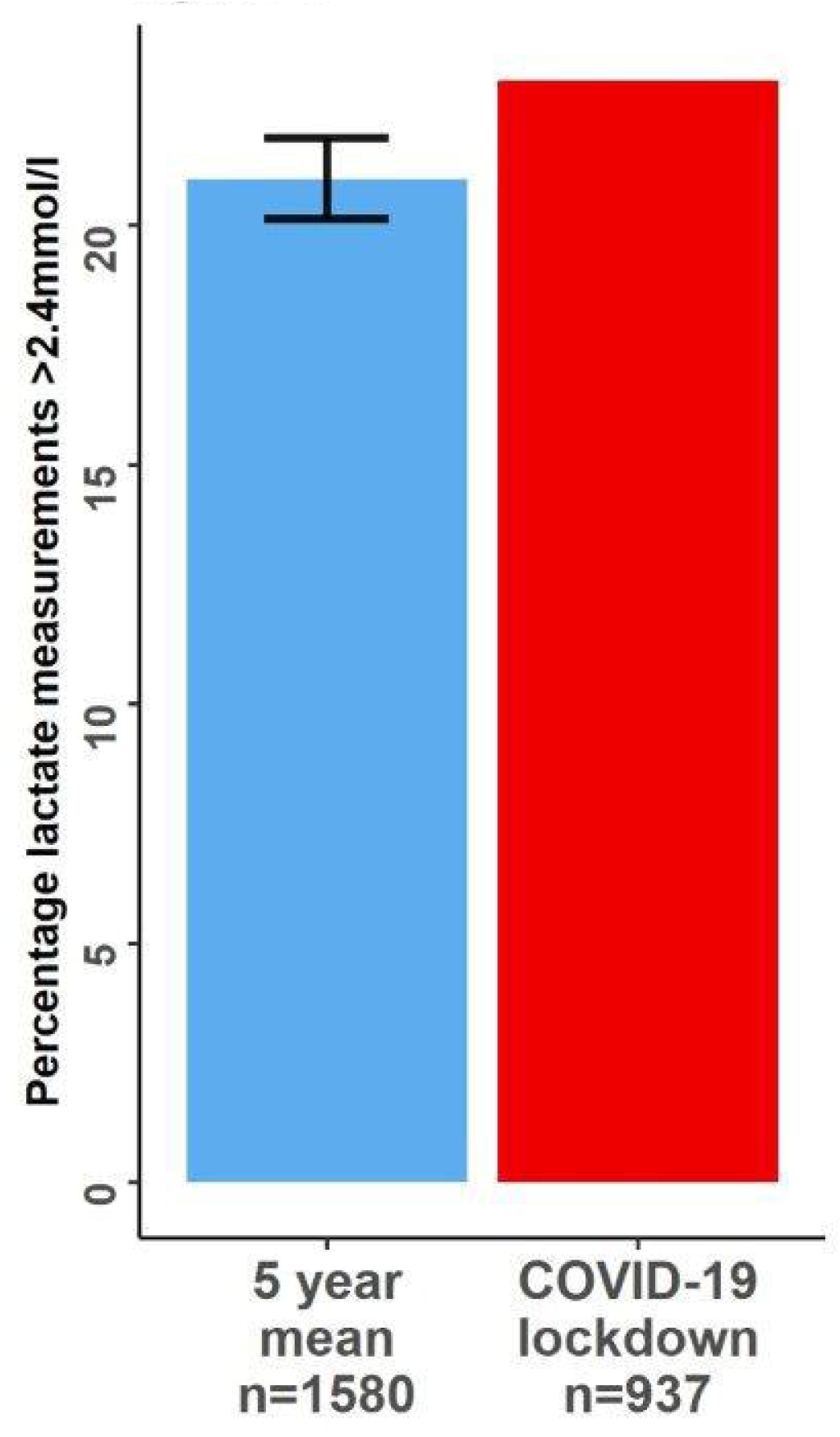
Proportion of patients with lactate above the reference range (2.4mmol/l) on admission. error bars = +/- standard error.

**Figure 6.**
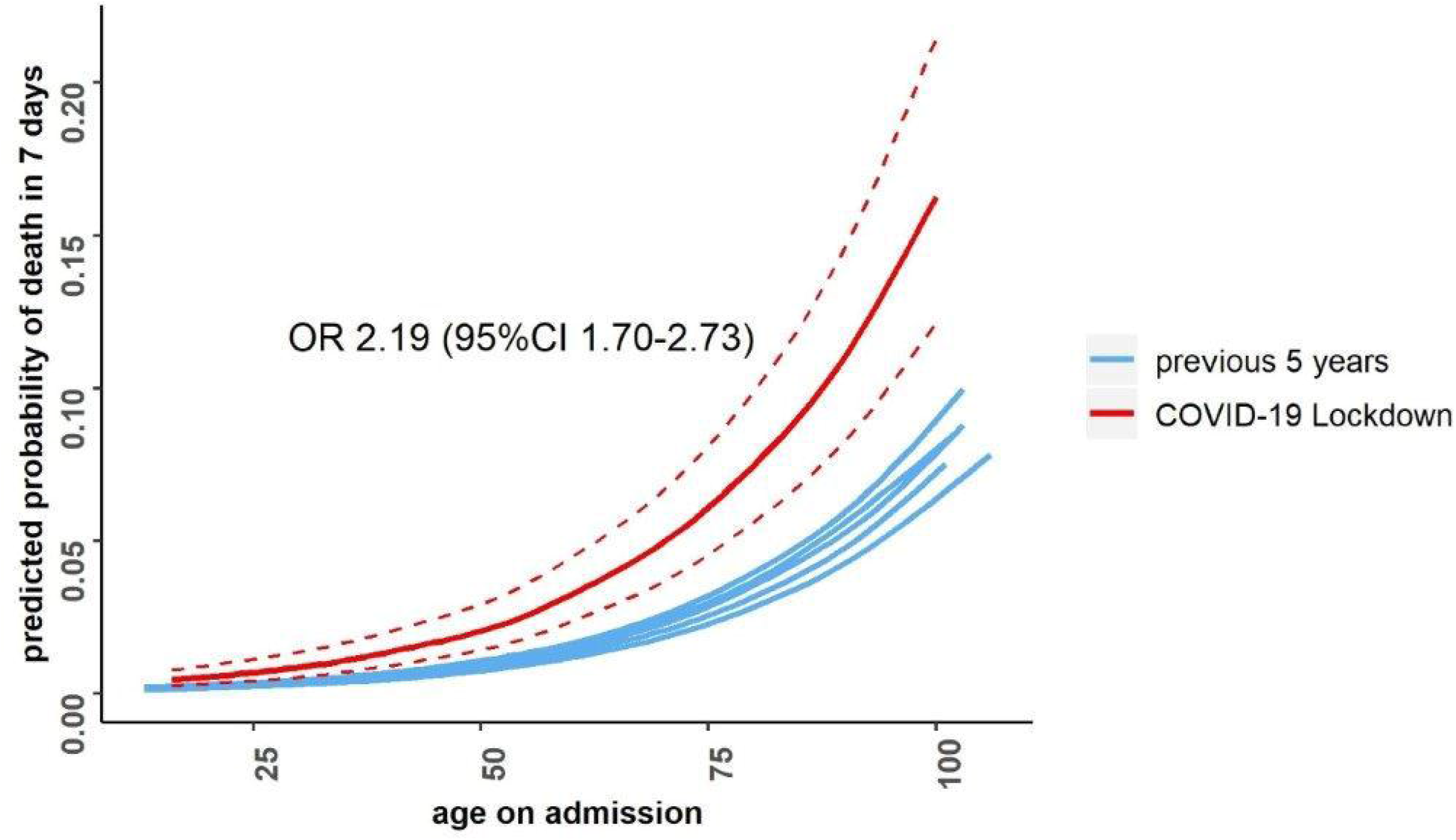
Binary logistic regression modelling of Age and SIMD adjusted 7 day hospital mortality of non-covid-19 admissions (n = 1776) versus 5 previous years (n= 14961). Dashed lines = 95%CI.

## Discussion

These data suggest that admissions to acute medical units have fallen considerably in the coronavirus pandemic and mirror observational studies in emergency care [2]. Alarmingly, patients admitted to AMUs are clinically more unwell with a higher incidence of acute kidney injury and lactic acidosis and a significantly higher incidence of in-hospital 7-day mortality.

There are several possible explanations for these findings. Late presentation with time sensitive pathologies such as sepsis, stroke or myocardial infarction worsens outcome [6–8]. It is possible that the higher level of illness acuity and higher mortality is due to this phenomenon.

This study includes patients with a lack of signs or symptoms of SARS-CoV-2 infection and those with features of COVID-19 in whom laboratory swab results were negative. The nose and throat swab test for SARS-CoV-2 is widely reported to have sensitivity limitations possible due predominance of the infection in the lungs with relatively little in the upper respiratory tract [9]. Locally we have found that 18% of patients admitted to hospital testing positive for SARS-CoV-2 were diagnosed on a subsequent follow up swab (unpublished data). Local guidelines which require repeat testing in those with a high clinical suspicion of COVID-19 may ameliorate this effect. Furthermore, patients with COVID-19 can present with atypical symptoms and signs, such as rash, seizures and gastrointestinal haemorrhage or stroke, which would not trigger testing under our local guidelines [10–12]. It may be therefore that a proportion of the increase in patient acuity and death observed is due to undiagnosed SARS-CoV-2 infection. The expansion of testing to include all hospital admissions in future may help to clarify this.

To the authors’ knowledge, this is the first study examining the non-COVID-19 acute medical admissions population during the pandemic. This use of fully electronic patient records with healthcare analytics allows patient level data to be collected quickly, accurately and prospectively and rapidly linked to laboratory and national public health datasets. Furthermore, the study population benefits from complete population coverage of all residents within a single administrative health region who were admitted.

There are limitations to this study. For expediency of reporting, disease coding and stratification of presenting pathology is not available at the time of writing and future analysis of case-mix and cause of death in this cohort may allow more detailed understanding of these findings. During the lockdown period, a higher proportion of patients with time-sensitive pathologies such as severe sepsis, myocardial infarction and stroke may bypass acute medical units and be directed to specialist units or critical care units. Furthermore, a higher proportion of patients may die in the community with similar pathologies. However, exclusion of these groups from the study population would bias our mortality findings towards the null.

## Data Availability

Subject to appropriate NHS Lothian governance approval we invite groups to submit requests for data sharing for the purposes of research and confirmatory analysis.

https://simd.scot/#/simd2016/

## Conclusion

The COVID-19 pandemic has been associated with a significant reduction in acute medical admission in all demographic groups. However, those attending are younger with greater medical acuity and a higher risk of inpatient mortality. Ongoing public health efforts must be made to ensure patients seek medical attention appropriately in the context of acute medical illness.

## Acknowledgements

Our sincerest thanks go to Stephen Young and Neil Murray and the team from Lothian Analytics Services at NHS Lothian for technical advice and expertise.

## Summary Box

### What is already known

Social distancing measures during the COVID-19 lockdown may be associated with non-covid-19 related excess morbidity and mortality due to a delay medical attendance. The effect of lockdown on presentation and outcomes in acute medical admissions without COVID-19 remains unknown.

### What this study adds

Non-Covid-19 acute medical admissions have reduced significantly during the lockdown period (46%) and are associated with a higher acuity at presentation and over 2-fold risk of early inpatient hospital mortality (OR 2.19).

Ongoing public health efforts must be made during the pandemic to ensure patients seek medical attention appropriately in the context of acute medical illness.

## Copyright

The Corresponding Author has the right to grant on behalf of all authors and does grant on behalf of all authors, an exclusive licence (or non exclusive for government employees) on a worldwide basis to the BMJ Publishing Group Ltd to permit this article (if accepted) to be published in BMJ editions and any other BMJPGL products and sublicences such use and exploit all subsidiary rights, as set out in our licence.”

## Contributors

Dr Marcus Lyall (Guarantor): Design of study, data collection and linkage, statistical analysis, manuscript preparation. Dr Nazir Lone: Study design, statistical analysis, and manuscript preparation.

## Ethics Approval

The study was reviewed by the Quality Improvement Team and registered in NHS Lothian as a Quality Improvement project. Following the HRA decision tool and after seeking advice from NHS Lothian Research and Development department, the study was deemed to be service evaluation and therefore formal ethical approval was not required. All data were anonymised before analysis and complied with local data protection requirements.

## PPI statement

We did not directly include PPI in this study, but the database used in the study was developed with PPI and is updated by a committee that includes patient representatives.

